# Personalization of Radiotherapy Dose in HPV Positive Oropharynx Cancer Using GARD

**DOI:** 10.1101/2023.09.14.23295538

**Authors:** Emily Ho, Loris De Cecco, Steven A. Eschrich, Stefano Cavalieri, Geoffrey Sedor, Frank Hoebers, Ruud H Brakenhoff, Kathrin Scheckenbach, Tito Poli, Kailin Yang, Jessica A. Scarborough, Shivani Nellore, Shauna Campbell, Neil Woody, Tim Chan, Jacob Miller, Natalie Silver, Shlomo Koyfman, James Bates, Jimmy J. Caudell, Michael W. Kattan, Lisa Licitra, Javier F. Torres-Roca, Jacob G. Scott

## Abstract

A central clinical goal for patients with HPV-positive oropharynx cancer has been to reduce radiation doses while maintaining cure rates. Recent results of Phase 3 prospective trial HN005 demonstrated that RT dose de-escalation can not be safely done based on clinical factors alone. We have previously shown that the genomic adjusted radiation dose (GARD) is predictive of radiation treatment benefit and can be used to guide RT dose selection. We hypothesize that GARD can be used to guide RT dose de-escalation in HPV-positive OPSCC patients.

Gene expression was analyzed for 191 formalin-fixed paraffin-embedded samples from HPV-positive OPSCC patients within an international, multi-institutional, prospective/retrospective observational study including patients with AJCC 8th edition stage I-III. Two RT dose fractionations were utilized for the majority of primary RT cases (70 Gy in 35 fractions or 69.96 Gy in 33 fractions). Median RT dose was 70 Gy (range 51.0-74.0), survival at 36 months and 60 months was 94.1% and 87.3%, respectively. Cox proportional hazards analyses were performed with GARD as a continuous variable and time-dependent ROC analyses compared the performance of GARD to clinical variables alone.

Despite near-uniform RT dosing, GARD reveals significant heterogeneity (range 15.4 - 71.7) in predicted effect. In univariate analysis, GARD was associated with an improvement in OS (HR = 0.941 (0.888, 0.998), p = 0.041). In multivariable analysis, each unit increase in GARD was associated with an improvement in OS (HR = 0.943 (0.891, 0.999), p = 0.046) where stage was not (T stage HR = 1.992 (0.711-5.576), p=0.190, N stage HR = 2.367 (0.867-6.460), p=0.093). ROC analysis for GARD at 36 months yielded an AUC of 78.26 (65.14, 91.38) compared with 71.20 (54.47, 87.93) for standard clinical variables. We identify two GARD-based strategies to RT dose personalization which are predicted to yield improved clinical outcomes, while delivering an average lower RT dose.

In this multi-institutional cohort of patients with HPV-positive OPSCC, GARD associates with OS, outperforms standard clinical variables and provides a novel genomic strategy to RT dose personalization. We propose that GARD should be incorporated in the diagnostic workup of HPV-positive OPSCC patients.

## INTRODUCTION

Since the discovery that human papillomavirus (HPV) is an etiologic and strong prognostic factor in oropharyngeal squamous cell carcinoma, assessing this biomarker indirectly via p16 or directly via *in situ* hybridization has become standard of care in the diagnostic and staging work up of these patients.^1,2^ Ang developed a three-group classification system based on clinical factors (HPV status, pack years of smoking, and T or N classification) that has informed the design of multiple clinical trials in the last decade.^3^ As the low risk group in this classification had an overall survival (OS) of 93% at three years, it was hypothesized that this favorable subset could be treated to a lower RT dose/toxicity without detriment in OS. Thus, the development of a successful uniform approach to RT dose de-escalation has been a central clinical aim of the field over the last decade.

While several approaches to RT dose de-intensification have been explored, all of them share a number of characteristics. First, they define eligibility based on clinical factors that define prognostic risk. For example, the NRG focused on low risk HPV+ patients receiving primary RT while the MSKCC approach^4^ focused on low risk patients with a negative F-MISO PET. Second, they assume all patients are biologically homogeneous and have the same opportunity to benefit from RT. Therefore, the RT de-escalation strategy utilized is uniform. NRG chose 60 Gy with concurrent cisplatin, while MSKCC chose uniform 30 Gy to the neck after surgery with no chemotherapy.

Recently, the NRG announced early results of HN005, a Phase 3 clinical trial testing the non-inferiority of uniform RT dose de-escalation (cisplatin + 60 Gy or nivolumab + 60 Gy) against the standard of care (cisplatin + 70 Gy).^5^ Unfortunately, the interim analysis failed to demonstrate the non-inferiority of cisplatin + 60 Gy over standard of care. These early results suggest that clinical factors and a uniform therapeutic approach are not enough to provide therapeutic guidance for RT de-intensification and suggest that similar to many targeted^6^ and immunotherapy agents,^7^ RT dose optimization may need to be targeted to specific genomically-defined subpopulations.

In previous studies, we developed the genomic adjusted radiation dose (GARD), a radiation-specific metric that quantifies the RT treatment effect in a given patient as a function of their RT dose and tumor genomics.^8^ GARD results suggest that the *treatment effect* of a uniform dose of RT (e.g. 70 Gy) is biologically highly heterogeneous, rather than homogenous, the current assumption in the field. In a recent pooled analysis of 1,615 patients in seven different disease sites, we demonstrated that GARD was associated with overall survival and recurrence risk as a continuous variable and predicted RT treatment benefit for each individual patient.^9^ Since GARD quantifies the treatment benefit for each individual patient, GARD-based models can be used to inform RT dose adjustments to optimize a patient’s clinical outome, thus providing a novel tool to personalize RT dose.

GARD provides a critical innovation compared with the current approaches to RT de-intesification: the ability to depart from the assumption that RT benefit is homogenous and the limitation of uniform RT dosing strategies. Since RT benefit is one of the critical factors defining clinical outcome in HPV-positive HNSCC patients, we hypothesized that GARD could provide novel information that will allow for better more personalized approaches to the successful treatment of these patients.

To test whether GARD could serve this purpose, we first assessed its prognostic ability in a modern cohort of radiation treated patients with HPV positive HNSCC. Further, we hypothesize that GARD’s prognostic information would provide an improvement to outcome prediction compared to clinical factors alone. Since GARD is intrinsically linked to radiation dose, any improvement in outcome prediction utilizing GARD can also fundamentally be used to make quantitative predictions for differential outcome given specific RT dose adjustments. This provides not just a tool for the definition of sub-populations for clinical trial design, but also the opportunity for truly personalized radiation dosing in the clinic.

Here we describe an analysis of patients treated with radiation therapy with HPV-positive HSNCC as part of the Big Data to Decide project.^10^ We use modern methods to assess individual patient radiation sensitivity indices (RSI)^11^ from gene expression data derived from formalin fixed tumor specimens, and use radiation-dosing information for each patient to calculate GARD. We then use continuous Cox proportional hazards regression to determine the relationship between GARD and outcome, and present a discrete analysis at several cutpoints *post hoc* to suggest optimal stratification strategies. Finally, we developed GARD-based models to personalize RT dose to achieve the best possible clinical outcome (both tumor and normal tissue) for each individual patient. We find that it is not only possible to reduce RT dose for a significant number of patients but that it is also possible to improve tumor outcomes with RT dose personalization.

## METHODS

### Patient Cohort

Patients in this analysis were part of the Big Data to Decide project (BD2Decide, NCT02832102), a collaboration of seven European centers to develop a clinico-genomic database of head and neck cancer patients. The details of this project have been previously described.^10^ Briefly, BD2Decide enrolled a total of 1,537 patients (1,086 retrospectively and 451 prospectively) with loco-regional advanced head and neck cancer (Stage III-IVa, IVb, AJCC 7th edition, or Stage I - III, AJCC 8th edition) treated with curative intent, including 377 patients with HPV-positive oropharyngeal cancer. Of these, 286 patients had gene expression profiling available (GEO data GSE163173).

As shown in **Supplemental Figure 1**, after excluding patients that were HPV DNA-negative (n=14), patients with locally-advanced disease that underwent single-modality treatment (n=37), patients treated with surgery alone (no GARD could be calculated) (n=1), and patients with post-operative RT (n=43), a final study population of 191 patients remained. The study was approved by institutional review boards of each of the participating institutions and when possible patients consented to enrollment or a waiver for consent was approved. Patients were treated between 2008 and 2017, and follow-up closed in September 2019. ^10^

HPV testing was performed with p16 immunohistochemistry and confirmed by HPV DNA testing following positive staining. ^10^ In total, 191 patients received definitive RT primary treatment. Fifteen RT dose fractionations were prescribed for definitive RT cases with the two most common being 70 Gy in 35 fractions or 69.96 Gy in 33 fractions. Median RT dose was 70 Gy (range 51-74) for definitive RT cases. The median follow up was 43.95 months (IQR 33.0-60.7).

### Bioinformatic and Statistical Analysis

All analysis was performed in R4.4.2 unless otherwise noted. Quarto markdown documents and data pre-processing scripts are available at https://github.com/steveneschrich/ `GARDHNC`. A full package list with versions is available as a ‘renv’ lockfile at the site above. Graphs were generated using ‘ggplot2’ and ‘ggpubr’ R packages.

All patient tumors previously underwent gene expression profiling using Affymetrix Clariom D from formalin fixed samples and are available from the Gene Expression Omnibus (GEO) as GSE163173. Raw CEL files were processed and normalized using Affymetrix sst-rma in Expression Console (Thermo Fisher), then RSI values were generated using a 10-gene signature as previously described,^12,13^ as implemented in the R package hacksig^14^. RSI has been previously clinically validated in multiple cohorts.^15–23^ A patient-specific genomic parameter, *α_g_*, was subsequently calculated using the linear-quadratic model to estimate patient radiosensitivity, the derivation of which we have previously described, yielding the relation:

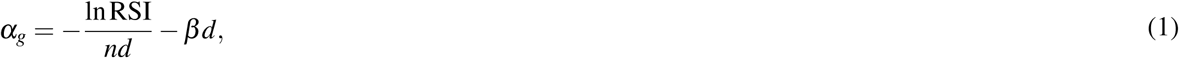

where dose *d* is 2 Gy, and the number of fractions, *n* is 1, as this moves from a genomic measure to the familiar Surviving Fraction after 2Gy (SF2). We further make the simplifying assumption that *β* is a constant at 0.05/Gy^2^. This genomic parameter, *α_g_*, is then used together with each patients specific radiation dose and fractionation to calculate their clinical GARD value (GARD*_c_*).

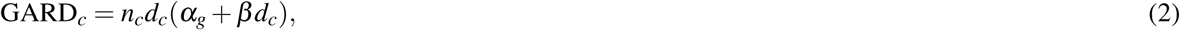

where *n_c_*is the number of fractions and *d_c_* is the dose per fraction per the clinically delivered radiation plan to each patient. These values were calculated without information about clinical outcome, using the described, previously specified model. Details of the derivation of *α_g_*and GARD*_c_* can be found in Scott et al.^24^.

Overall survival comparisons were performed using the ‘survival’, ‘gtsummary’ and ‘rms’ packages. Cox proportional hazards regression from the R package ‘rms’ was used to assess the association between GARD as a continuous variable and overall survival (OS). Multivariable Cox regression analysis was performed using ‘survival’ and ‘gtsummary’ packages. Survival curves were generated using the R package ‘survminer’. Discrete analyses with log-rank statistics were performed for hypothesis generation, using an algorithm to minimize the logrank statistic to derive optimal groups in two dose levels. The R packages ‘survminer’ and ‘maxstat’ were used for this analysis.

OS was defined as the time between primary diagnosis and death or last follow-up. Follow up was censored at 60 months if no event occurred prior. After performing standard analyses to determine the association of GARD with outcome, we incorporated GARD with known prognostic clinical variables including stage, pack-years smoking and ECOG performance status to determine whether GARD improved prognostic performance. In addition, we also integrated the three-cluster gene expression model^12^ following standard methods.^25^ The Cox proportional hazard models generated by the ‘rms’ R package were evaluated by comparison of time-dependent receiver operating characteristic (ROC) curve analysis with the `timeROC` package in R.^26^

### *In silico* clinical trial simulations

We first performed an *in silico* trial to test uniform RT dose de-intensification (from 70Gy to 60Gy) in this cohort of HPV-positive oropharyngeal cancer patients. Virtual patients were generated by sampling from the density estimates of the empirical RSI distribution from the BD2DECIDE cohort. Outcomes are estimated based on a Weibull distribution of the original outcomes evaluated at discrete timepoints. The combination of low/high GARD population distributions yield the overall cohort outcome estimates. GARD was calculated for each virtual patient, and an OS curve was predicted based on the GARD level achieved, using the optimized two dose GARD level approach (GARD *<* 42 = low and GARD *≥* 42 = high). This cutpoint was identified as previously described, see **Supplemental Figure 4**.

To determine whether GARD could identify a successful de-intensification strategy, we performed a two arm *in silico* trial in which only a GARD-identified subset of patients were de-intensified in one arm. The control arm was treated uniformly at 70 Gy, as above. In the *in silico* experimental arm, only GARD-high patients (GARD *≥* 42) who remained in the same risk group after de-escalation to 60 Gy were de-intensified. All other patients remained at 70 Gy. In other words, GARD-low (high-risk) patients are treated at 70 Gy, and GARD-high patients are de-intensified to 60 Gy unless this changes their risk category.

As a final, exploratory analysis, we sought to determine what a purely personalized approach, with a GARD target chosen to provide an overall outcome equivalent to our current standard of care, would reveal. In this personalized iso-curative dosing strategy, we compared a standard of care arm (70 Gy in 35 fx) to a target GARD identified to provide equipoise to modern outcomes (in this case GARD = 32), see **Supplemental Figure 5**. We then calculated the total difference in dose predicted to provide equipoise across the population, and the differential in cost to provide this at the population level.

## RESULTS

### HPV-positive oropharyngeal squamous cell carcinoma cohort

We identified 191 patient tumors previously profiled through the BD2DECIDE project meeting the criteria of definitive RT^12^ (see **Supplemental Figure 1** for details on patient selection). The characteristics for these HPV-positive oropharyngeal squamous cell carcinoma patients are detailed in **Supplemental Table 1**.

### GARD reveals underlying genomic heterogeneity in RT effect

We have previously shown that GARD reveals underlying heterogeneity in radiation treatment effect within groups presumed to have been treated uniformly (with approximately equivalent physical dose).^8,9,19,20^ In this cohort we demonstrate again that GARD reveals wide heterogeneity in predicted RT effect in spite of relatively uniform RT dose prescribed. As shown in **Figure 1 (Left)**, delivered GARD ranged from 15.4 to 71.7 (median: 39.1, IQR: 12.6). Plotted along the edges of the jointplot between GARD and EQD2 are kernel density estimates for the entire cohort, revealing wide heterogeneity in delivered GARD (IQR 12.6) in the setting of near homogeneity in RT dose (IQR 0.04). The difference between GARD and EQD2 is best exemplified by the patients who received the whole course of ‘standard’ radiation dose – with EQD2 measures between 69-71 Gy, see **Figure 1 (Right)**. The range of GARD for those patients was 19.7-71.7 (IQR 12.7) even though they all were treated to (approximately) the same RT dose (EQD2), highlighting the wide differential in the predicted effect of our uniform clinical dosing strategies. The distributions of RSI and GARD by AJCC8 stage did not differ significantly (**Supplemental Figure 2** and **Supplemental Table 2**).

**Figure 1.**
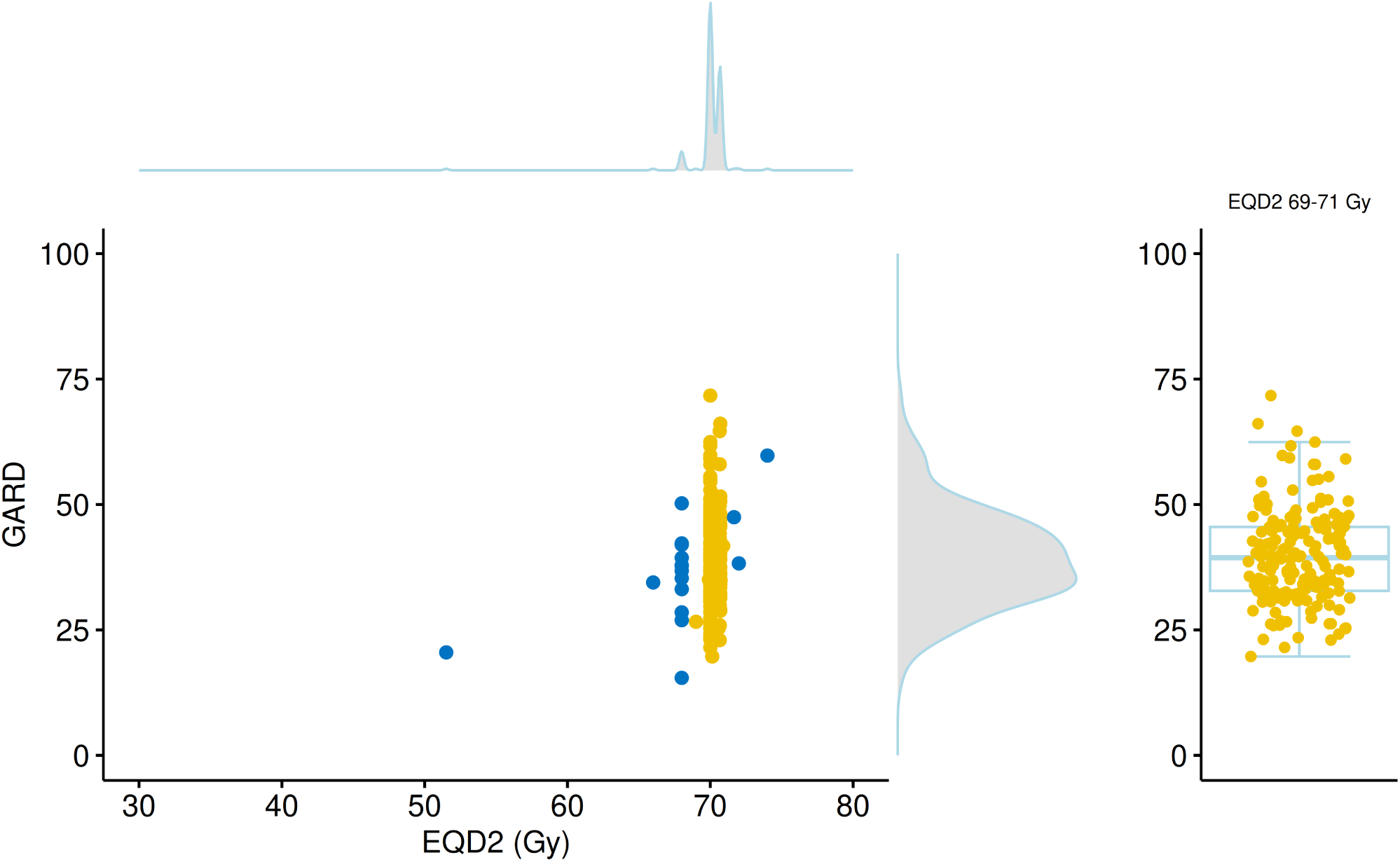
GARD exhibits large underlying genomic heterogeneity in radiation effect compared to radiation dose alone. Left: EQD2 (median 70.0, IQR 0.7) is plotted against associated GARD (median 39.1, IQR 12.6) for each patient in the cohort. Kernel density estimates are plotted on each edge to show the distributions of the individual variables. Patients that received an EQD2 of 69-71 Gy (standard dosing) are indicated in yellow color. **Right:** The GARD value of all patients receiving an EQD2 of 69-71 Gy (standard of care) highlight GARD’s ability to stratify patients by their genomic heterogeneity. Data points are overlaid with a box-whisker plot with box representing quartiles and whiskers extending to 1.5 times IQR.

### GARD is continuously associated with OS in RT-treated HPV-positive OPSCC patients

Previously, we demonstrated that GARD was associated with overall survival, recurrence risk and was predictive of RT benefit in a pooled pan-cancer analysis including 1,615 patients which included cohorts with HSNCC.^9^ Since RT therapeutic benefit is a critical factor impacting clinical outcome in HPV-positive patients, we hypothesized that GARD would be associated with clinical outcome in this analysis of HPV-positive oropharyngeal squamous cell carcinoma patients collected through the BD2DECIDE project.^12^ To test this, we performed a Cox proportional hazards analysis of GARD and OS in patients that were treated with definitive primary RT (n=191) and those treated with standard of care definitive primary RT (EQD2 69-71Gy) (n=174). As shown in **Figure 2, Left**, GARD is associated with OS as a continuous variable for patients treated with primary definitive RT and censored at 60 months. We found that for each unit increase in GARD there is an improvement in OS (HR (95% CI) = 0.941 (0.888, 0.998) *per unit* GARD, p = 0.041). This association of GARD with OS also held when including only patients treated with primary definitive RT at standard of care doses (EQD2 69-71Gy) as shown in **Figure 2, Right** (HR (95% CI) = 0.920 (0.857, 0.986) per unit GARD, p = 0.019). This suggests that GARD can stratify patients by predicted *effect* even when radiation dose is approximately uniform.

**Figure 2.**
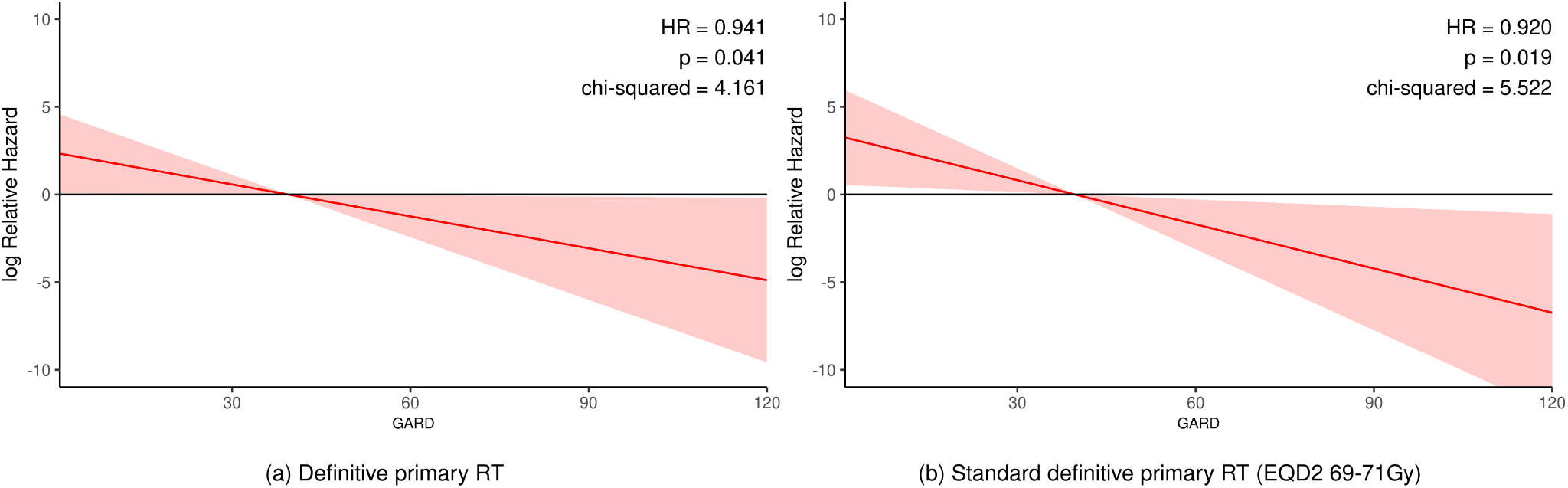
GARD is a continuous predictor of OS in radiation treated patients with HPV-positive OPSCC. **(a)** Cox proportional hazards analysis demonstrates significant continuous association between GARD and OS for patients treated with primary definitive RT and censored at 60 months (p = 0.041, HR = 0.941 (0.888, 0.998) *per unit* GARD). **(b)** Cox proportional hazards analysis demonstrates significant continuous association between GARD and OS for the subset of patients treated with standard of care primary definitive RT (EQD2 69-71Gy) (p = 0.019, HR = 0.920 (0.857, 0.986) *per unit* GARD).

### GARD is associated with OS in RT-treated HPV-positive OPSCC patients

The BD2DECIDE cohort includes clinical variables for performance status (ECOG > 0), T stage (T4 vs T1-3), N stage (N2-N3 vs N0-N1), and smoking pack years (>10). We performed multivariable analysis using these variables and GARD for statistical associations with OS. In the definitive primary RT cohort, GARD was the only variable statistically associated with OS (HR = 0.943 (0.891, 0.999), p = 0.046). These results are summarized in **Table 1**.

**Table 1.** Multivariable analysis of definitive primary RT patients. GARD is associated with overall survival, censored at 60 months (p = 0.046).

|  | HR (95% CI) | p |
| --- | --- | --- |
| **GARD (continuous)** | 0.943 (0.891, 0.999) | **0.046** |
| **T stage (T4)** | 1.992 (0.711, 5.576) | 0.190 |
| **N stage (N2-N3)** | 2.367 (0.867, 6.460) | 0.093 |
| **ECOG = 1 or 2** | 0.908 (0.247, 3.342) | 0.884 |
| **Pack-years (>10)** | 2.117 (0.756, 5.929) | 0.154 |

In addition, we developed and evaluated a Cox regression model including the previously known prognostic clinical variables (T stage, N stage, smoking and ECOG performance status), to determine whether a model including GARD improves overall model performance. As shown in **Figure 3**, the Cox model including clinical variables achieves an AUC of 71.20 whereas GARD alone achieves a superior AUC (3 yr OS) of 78.26. Integrating GARD with the clinical variables improves the prognostic ability of the model with AUC 83.81. We also evaluated the previously developed 3-cluster model^12^ which, by itself, achieves an AUC similar to the clinical variable model (AUC: 72.83). However, integration of the 3-cluster information into the GARD + clinical variable model does not improve the overall prognostic ability as measured by AUC 83.81. This lack of improvement may be related to the relationship between clusters and GARD values (see **Supplemental Figure 3**).

**Figure 3.**
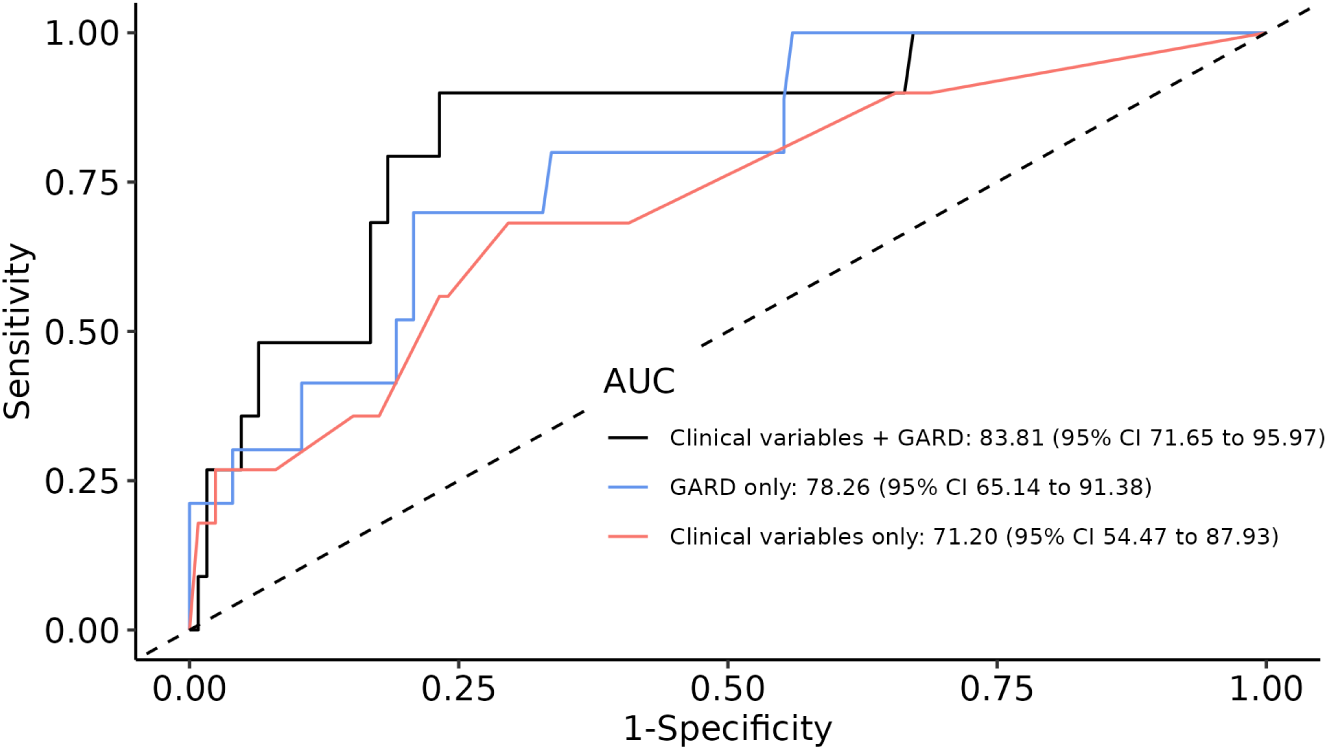
AUC analysis shows GARD outperforms standard clinical variables. In this AUC analysis we compare Cox regression of clinical variables (red) to GARD alone (blue) and a combined model using all factors (black). The combined model shows dramatic improvement compared to clinical variables alone. This model included all 191 definitive primary RT patients, and analysed outcome at 3 years.

### GARD predicts that empiric dose de-escalation would result in inferior clinical outcome

Although HPV- positive patients have excellent prognosis, the interim analyses of HN005 have emphasized the importance of developing clinical tools to identify patient subsets with differential risk of clinical failure. We hypothesized that GARD could identify a sub-population of HPV-positive patients at differential risk of failure that may explain the failure of unselected empiric dose de-escalation as tested in HN005. In addition, understanding the differential risks of failure can lead to a better clinical strategy for dose de-escalation in selected patients. To develop this, we performed an exploratory discrete analysis based on an optimized cut-point analysis. Minimizing the log-rank score at one discrete value reveals two groups with maximally different outcomes (see **Supplemental Figure 4**). This analysis revealed one cutpoint at GARD *<* 42 which optimally stratified patients as shown in **Figure 4A**.

**Figure 4.**
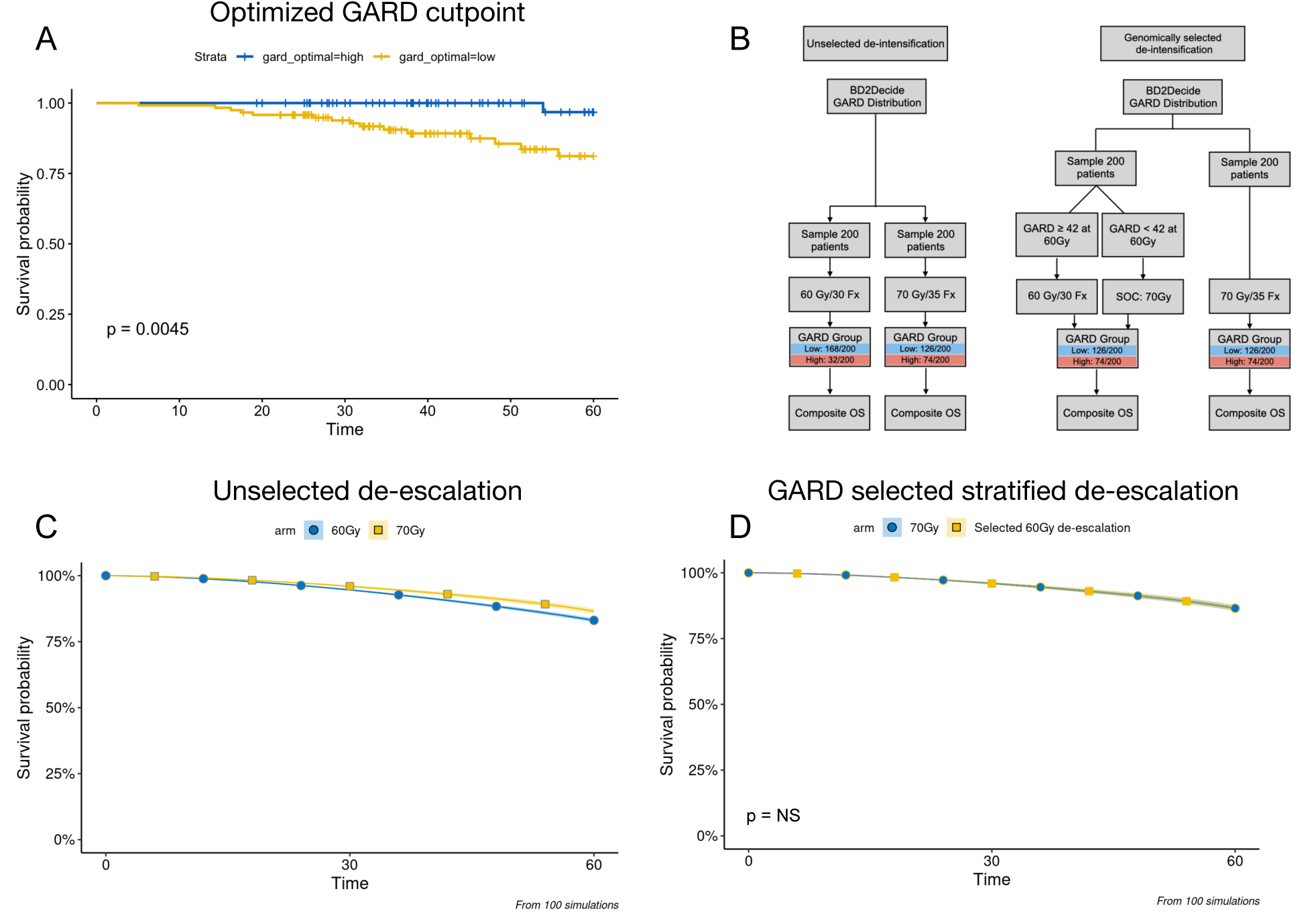
GARD predicts that uniform RT dose de-escalation in HPV-positive patients would result in an inferior clinical outcome compared to standard of care. **(A)** GARD identifies HPV-positive patient subsets with differential risk of failure. An exploratory analysis identified one cut-point which group patients in two risk levels. Patients that achieve the lowest GARD (< 42) have a higher risk of failure (3 year OS = 90.5%). (B) *In silico* clinical trial designs. **Left:** We simulate unselected RT dose de-escalation (cisplatin + 60 Gy vs cisplatin + 70 Gy). We utilize the RSI distribution of the BD2Decide cohort to generate GARD for 400 virtual patients randomized to either 60 or 70 Gy, and repeat this 100 times. **Right:** GARD-based De-escalation. In one example of a potential trial, we simulate a GARD-selected trial where only patients with GARD *≥* 42 are eligible for randomization to de-intensification. **(C)** Simulation of unselected RT dose de-escalation (cisplatin + 60 Gy), results in inferior OS compared to standard of care (cisplatin + 70 Gy). The unselected *in silico* clinical trial predicts that patients treated with RT dose de-escalation experience a statistically significantly worse overall survival when compared to standard of care (3-yr OS 92.7% vs 94.6%, non-overlapping confidence intervals). **(D)** Selective de-intensification produces similar OS.

Patients that achieved the GARD high (GARD *≥* 42) had a 3yr-OS of 100% (CI: 1-1) compared with 90% (CI: 0.85-0.96) for the GARD low group (GARD *<* 42). These differences are statistically significant with p = 0.0045, though this analysis should be interpreted carefully as it was performed for hypothesis generation and the groups were chosen by maximizing differences *post hoc*.

One possible explanation for the HN005 results is that empiric dose de-escalation results in a small number of patients falling from the GARD high cohort (GARD *≥* 42) to the GARD low cohort (GARD *<* 42) leading to an inferior result for empiric dose de-escalation. To test this hypothesis, we performed an *in silico* clinical trial to evaluate GARD-based predictions of clinical outcome for empiric dose de-escalation to 60 Gy (with concurrent chemotherapy) as in HN005. We found that GARD predicts that empiric (unselected) dose de-escalation would result in an inferior clinical outcome. The predicted 3 yr OS for patients modeled at 70 Gy is 94.6% compared with 92.7% for patients modeled at 60 Gy (**Figure 4C**). Empiric, unselected dose de-intensification is predicted to increase the proportion of patients in the GARD low group while decreasing the proportion of patients in the GARD high group. The 70Gy *in silico* arm had an average of 126 and 74 patients in the low and high GARD groups, while the 60Gy *in silico* arm had 168 and 32 patients in those groups.

Next, we determined whether we could use GARD to develop a clinical trial strategy that would predict equivalent outcome at 70 or 60 Gy. In one approach, GARD can identify patients that would remain at or above the GARD-high cutpoint (*≥* 42) at 70 or 60 Gy. Based on simulations, approximately 16% of the HPV-positive trial population would be eligible for dose de-escalation in this scenario. It should be noted that this approach excludes GARD-low patients and patients that fall from GARD-high to GARD-low at 60 Gy. The predicted OS curve for this approach to de-escalation is shown in **Figure 4D**. The 36 month survival proportion is equivalent (94.6%) in both arms of this simulated trial.

Another way to think about dose de-escalation is to ask the question: “can GARD identify a personalized target dose with the goal of *maintaining current outcomes*?” This is fundamentally different than our previous approach which asked if we could use GARD to select patients for stratified de-escalation to standard dose levels. In this approach we instead ask what GARD cutpoint would provide equipoise to current standard of care? Analyzing our cohort through this lens, we find that a significantly lower GARD cutpoint equal or higher than 32 (see **Supplememntal Figure 5**) would provide outcomes in line with current standard of care. **Fig 5A** shows the outcome of patients that achieved GARD 32 compared to unselected patients in the BD2Decide cohort. As shown, the patients that achieve a GARD of at least 32 have the same OS as the whole unselected cohort, thus achieving equipoise with current SOC in unselected patients.

**Figure 5.**
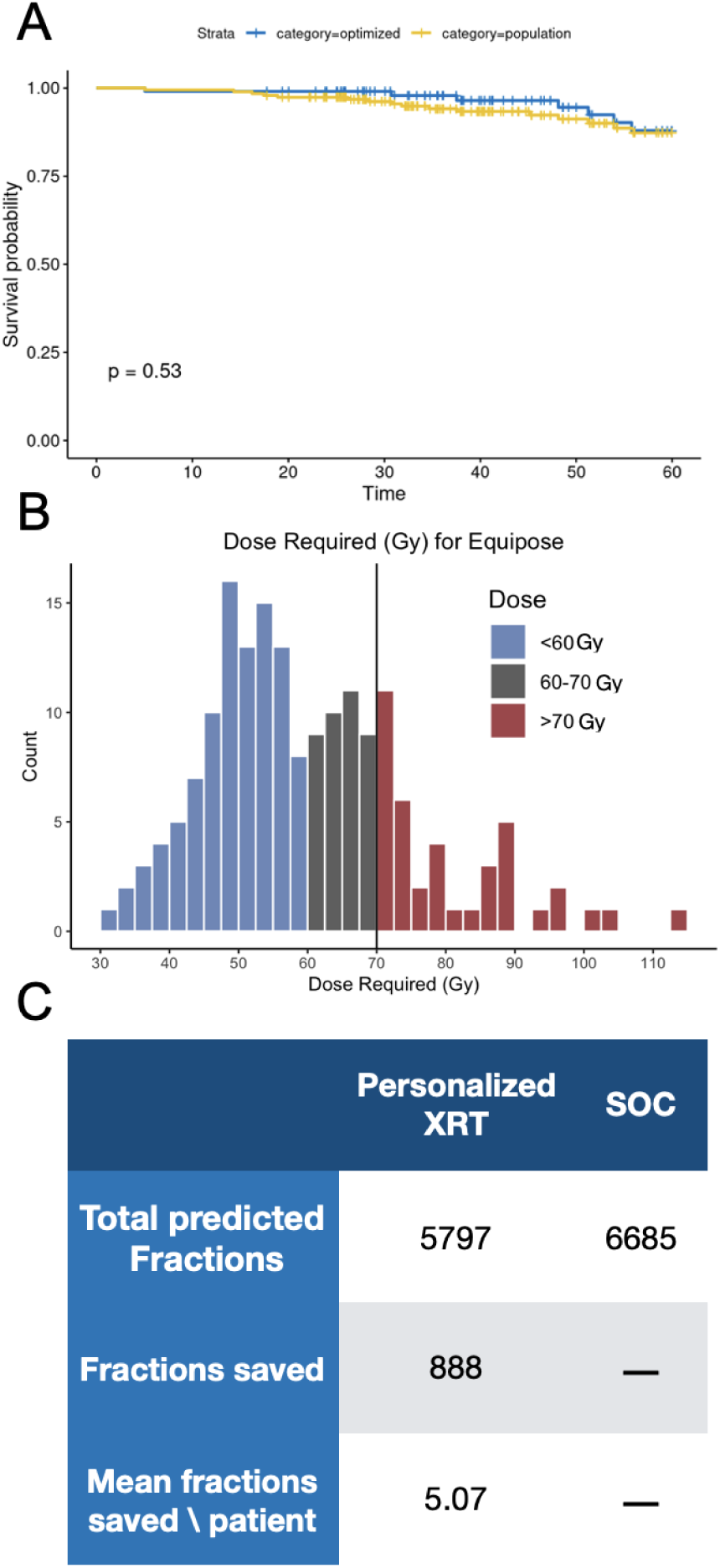
GARD-based opportunities for equipoise targeted RT dose reduction: maintaining equivalent outcomes to standard of care. **(A)** GARD targeted equipoise RT dose reduction. KM curves show that patients in the BD2Decide cohort that achieve a GARD of at least 32 achieve isocurative outcomes compared with the unselected cohort. Each patient can then have a prescription RT dose to match the target GARD (at least 32). Comparing this to SOC provides equivalent outcomes. **(B)** A histogram depicting the difference between the dose predicted to be required for each patient compared to the dose delivered. Red suggests patients were underdosed (39/175), offering opportunities to increase oncologic outcomes, and blue overdosed, indicating opportunities to decrease toxicity. **(C)** Calculating the difference between fractions delivered in SOC compared to the number predicted on a per patient basis reveals a large potential for toxicity reduction at the population level. This averages approximately 5 fractions (one week of radiotherapy) per patient, but with large heterogeneity.

In a trial designed like this then, each patient would be assessed for their RSI, and then a physical radiation dose would be calculated such that they would achieve a *prescribed GARD* of at least 32. While exploratory and non-standard, this analysis of a *genomic prescription paradigm* offers us a window into the future where dose is truly personalized – allowing exactly enough radiation to be delivered for tumor cure, minimizing toxicity. In **Fig 5B** we calculate the minimum dose required for each patient in the BD2Decide cohort to achieve a GARD of at least 32. Interestingly, the average dose needed aligns well with our clinical intuition – approximately 60Gy – but with large heterogeneity across individual patients. Of note as well is the large number of patients (22.3 percent in this cohort, 39 / 175) who we predict require between 60 and 70 Gy, revealing which patients would have inferior outcomes when de-escalated to 60 Gy.

This also suggests that on average the toxicity (financial and clinical), of nearly 5 fractions/patient can be spared while maintaining similar outcomes (**Figure 5C**). However the potential toxicity reduction for each patient is variable with some patients predicted to only require 30 Gy while a small minority may require higher doses than standard. Of note, there is a peak in the distribution between 60 and 70Gy, meaning that as we reduce dose from 70 to 60Gy without genomic guidance, we underdose a significant portion of patients, worsening our outcomes. This stands in contrast to our findings in non-small cell lung cancer,^24^ where the *dose escalation* from 60 to 74Gy (as in RTOG 0617) spanned a valley in the distribution, meaning that the escalation resulted in very few patients being benefited, while all received the increased toxicity.

## Discussion

The development of prognostic and predictive models to more accurately prescribe therapies is a central goal of personalized oncology. In this paper, we show that GARD, a previously described predictive model of the treatment effect of RT, is associated with overall survival in HPV-positive oropharyngeal cancer patients treated with RT as a continuous and optimized dichotomous variable. Furthermore, using time-dependent ROC analysis, we show that GARD outperforms standard clinical variables for prediction of OS of these patients, and that combining genomics (GARD) together with clinical factors provides a superior model to either alone.^27^ Finally, we show that GARD provides an opportunity to depart from the uniform RT dosing strategies that are hindering the improvement in outcomes for our patients. Instead of searching iteratively for subgroups that can be uniformly dose-optimized, we propose a strategy where each patient can have their dose (and plan) optimized based on their individual biology.

With the aim of breaking the mold in radiation oncology trial design, we propose two different GARD-based strategies to treatment optimization. The first prioritizes individual oncologic outcome for every patient, without considering concomitant toxicities at the population level. We find that personalizing RT dose to achieve GARD *≥* 42 maximizes oncologic outcome in this dataset. However, only 16 percent of patients achieve GARD *≥* 42 at 60 Gy, limiting this approach if the goal is to reduce toxicity. In the second approach, we define a risk of failure that matches the risk of an unselected population treated at 70 Gy. This allows us a larger opportunity to reduce RT dose and toxicity risk. If we based the GARD model on these parameters, we find that personalizing RT dose to achieve GARD of at least 32 achieves equipoise with current population level oncologic outcomes while reducing RT dose, and therefore toxicity, to the majority of patients.

In this approach a larger proportion of patients become candidates for de-escalation (77.7 percent), while a small fraction are identified for potential escalation (22.3 percent). This strategy matches current clinical outcomes but delivers an average of 5 fewer fractions per patient and thus achieves the intended aim of HN005 of equipoise at an average dose of 60 Gy. However the key difference is that the personalized dose for each patient is not 60 Gy, but instead, like most polygenic biological traits, a wide range (between 31-113 Gy). While the majority of patients require lower doses a small minority may need dose intensification or more a more effective radiosensitization strategy with concurrent chemotherapy. Further work on chemotherapy sensitivity^28^ and balancing toxicity with tumor control^24^ could also inform future iterations of these trials, most of which include cisplatin as a radiosensitizer.^29^

In a previous study, we introduced the concept of the toxicity cost of RT dose personalization; the optimization of RT dose can result in increased or reduced risk of normal tissue toxicity.^24^ And this toxicity cost/gain is also different for each patient. In this analysis, GARD proposes a genomic-based strategy that achieves clinical equipoise while decreasing average dose by 5 fractions/patient. Thus by accounting for biological heterogeneity, GARD-based RT prescription provides critical data that may improve the therapeutic ratio of RT; maximizing clinical outcome at the lowest possible toxicity risk.

In conclusion, we demonstrate that GARD outperforms standard clinical variables as a prognostic biomarker in HPV-positive OPSCC patients and provides a biology-based approach to radiotherapy dose personalization. GARD predicts that while the majority of patients require lower doses than SOC, a small minority of resistant patients still require 70Gy or potentially a higher dose to maintain their excellent clinical outcome. Thus an unselected uniform RT dose reduction approach fails because it does not recognize biological heterogeneity in radiosensitivity. To fully realize the clinical potential of radiotherapy in HPV-positive Oropharynx cancer patients, we must integrate the assessment of biological heterogeneity into our treatment strategy. Integrating GARD into the clinical evaluation of HPV-positive oropharynx patients is a step in that direction – bringing radiation oncology forward one more step towards truly personalized medicine.

## Data Availability

All data and code produced in the present study are available upon reasonable request to the authors or in the associate github.

https://github.com/steveneschrich/GARDHNC

## Supplemental Information

### Code and data availability

Statistical analyses were conducted using R v4.4.2 and associated packages. All quarto/R scripts and data pre- processing R scripts are available at https://github.com/steveneschrich/GARDHNC.

### Study Cohort

Patients from this analysis were part of the BD2Decide project. HPV-positive oropharyngeal cancer patients (N=234) with primary definitive RT and evaluable RSI/GARD (**Supplemental Figure 1**) were identified and characterized (**Supplemental Table 1**).

**Supplemental Figure 1.**
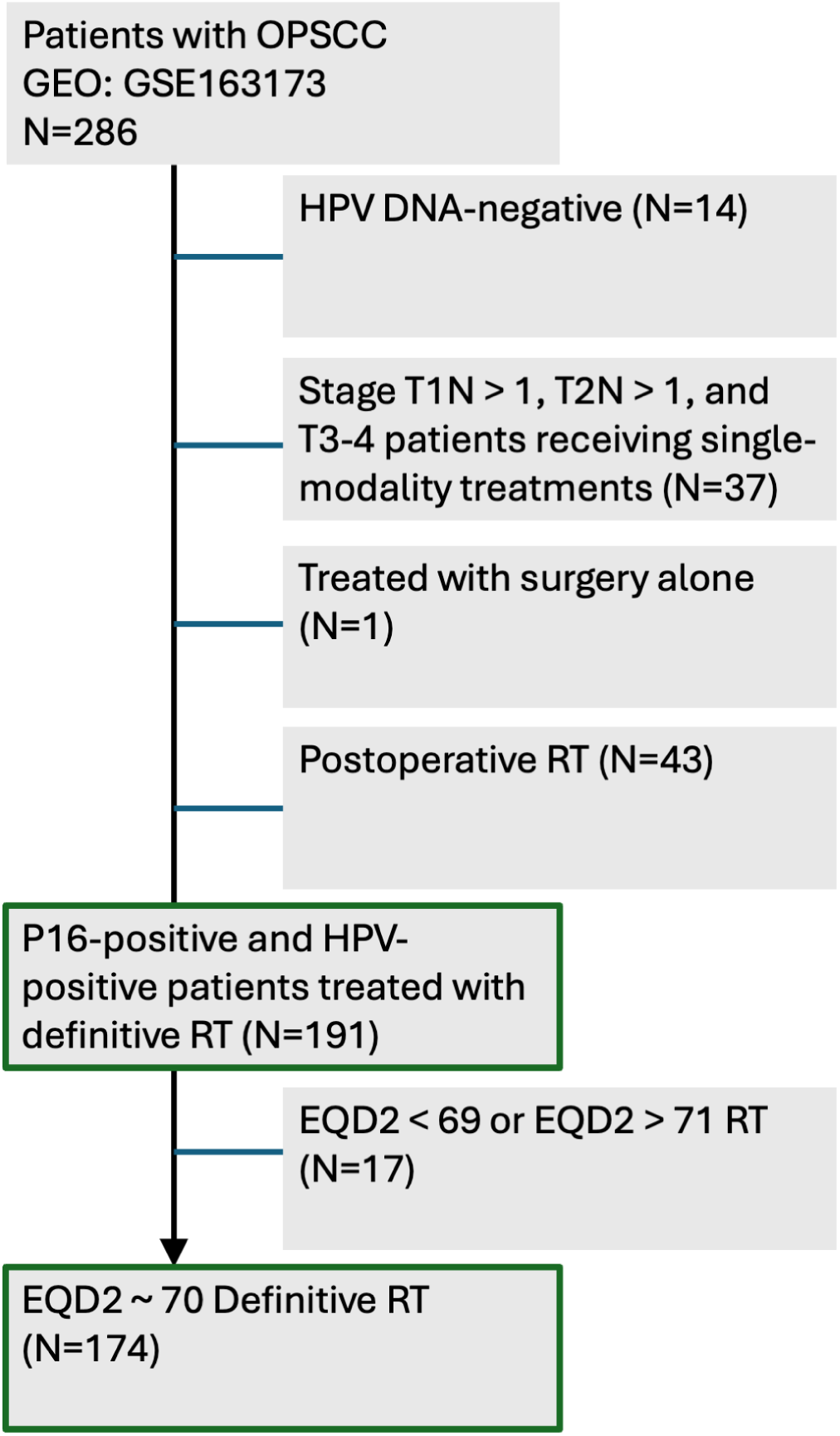
CONSORT diagram for study cohort. Green boxes indicate cohorts used within this work.

**Supplemental Table 1.** Characteristics of identified BD2DECIDE patient cohort (N=191). Counts are provided in parentheses except for Smoking Pack Years, which is reported as median and interquartile range (IQR).

| Characteristic | N = 191 |
| --- | --- |
| AJCC8, n (%) |  |
| I | 87 (46) |
| II | 49 (26) |
| III | 55 (29) |
| ECOG, n (%) |  |
| 0 | 158 (83) |
| 1 | 32 (17) |
| 2 | 1 (0.5) |
| Smoking Pack Years, Median (IQR) | 8 (0 – 30) |
| AJCC8 T stage, n (%) |  |
| T1 | 38 (20) |
| T2 | 57 (30) |
| T3 | 45 (24) |
| T4 | 51 (27) |
| AJCC8 N stage, n (%) |  |
| N0 | 6 (3.1) |
| N1 | 143 (75) |
| N2 | 34 (18) |
| N3 | 8 (4.2) |
| status, n (%) |  |
| Alive | 172 (90) |
| Dead | 19 (9.9) |

### RSI and GARD Distributions

Distributions of RSI and GARD grouped by AJCC8 stage are shown in **Supplemental Figure 2**.

**Supplemental Figure 2.**
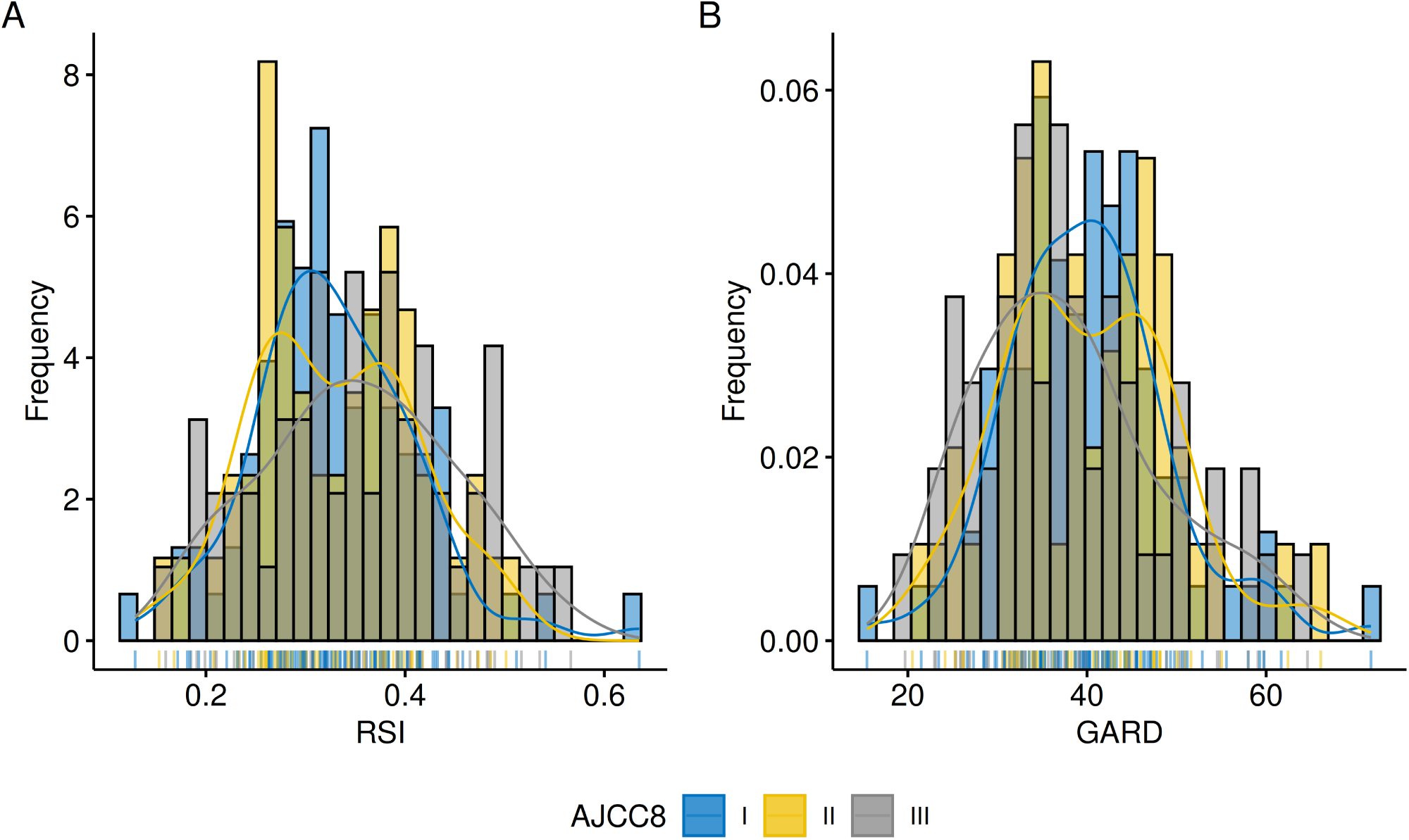
The distributions of RSI and GARD for each AJCC8 stage are similar.

Kruskal-Wallis tests revealed no significant differences among these distributions (by stage), as shown in **Supplemental Table 2.**

**Supplemental Table 2.** The p-values from the Krusal-Wallis rank sum test for differences in RSI/GARD distributions across stage. Median (IQR, interquartile range) values shown.

|  | Stage | N | Median (IQR) | p |
| --- | --- | --- | --- | --- |
| RSI | I | 87 | 0.32 (0.28 – 0.38) | 0.23 |
|  | II | 49 | 0.33 (0.26 – 0.39) |  |
|  | III | 55 | 0.35 (0.28 – 0.41) |  |
| GARD | I | 87 | 40 (34 – 46) | 0.25 |
|  | II | 49 | 39 (34 – 47) |  |
|  | III | 55 | 37 (31 – 45) |  |

### AUCs for various predictors

Comparison of the 3 most significant predictors individually via ROC analysis showed the greatest AUC for GARD alone (78.26), while the model combining GARD, T stage, N stage, smoking and ECOG status produced the highest AUC (83.81); values are listed in **Supplemental Table 3**. Of note, if RSI is compared here with the same cohort, a similar score to GARD is achieved of 77.68 (95% CI: 65.13 to 90.23) as the dose range in this cohort is narrow.

**Supplemental Table 3.** The time-dependent AUC and 95% CI at 3 years are shown for the predictors individually as well as for the combined nomogram.

|  | 3-yr AUC (95% CI) |
| --- | --- |
| **Clinical** | 71.20 (54.47, 87.93) |
| **Clusters** | 72.83 (59.01, 86.65) |
| **GARD** | 78.26 (65.14, 91.38) |
| **Clinical + GARD** | 83.81 (71.65, 95.97) |
| **Clinical + GARD + Clusters** | 83.81 (71.65, 95.97) |

### GARD vs Clusters

The prognostic gene expression clusters defined by Locati^30^ and Cavalieri^12^ are related to the GARD levels (particularly for patients with low GARD/High Risk), see **Supplemental Figure 3**. Kruskal-Wallis tests revealed significant differences in GARD among these clusters. Note that in the original publications the clusters were reported as RSI distributions, which we have transformed here into GARD using **Eq 1** and **2**. We have kept the cluster numbers as per the original publication.

**Supplemental Figure 3.**
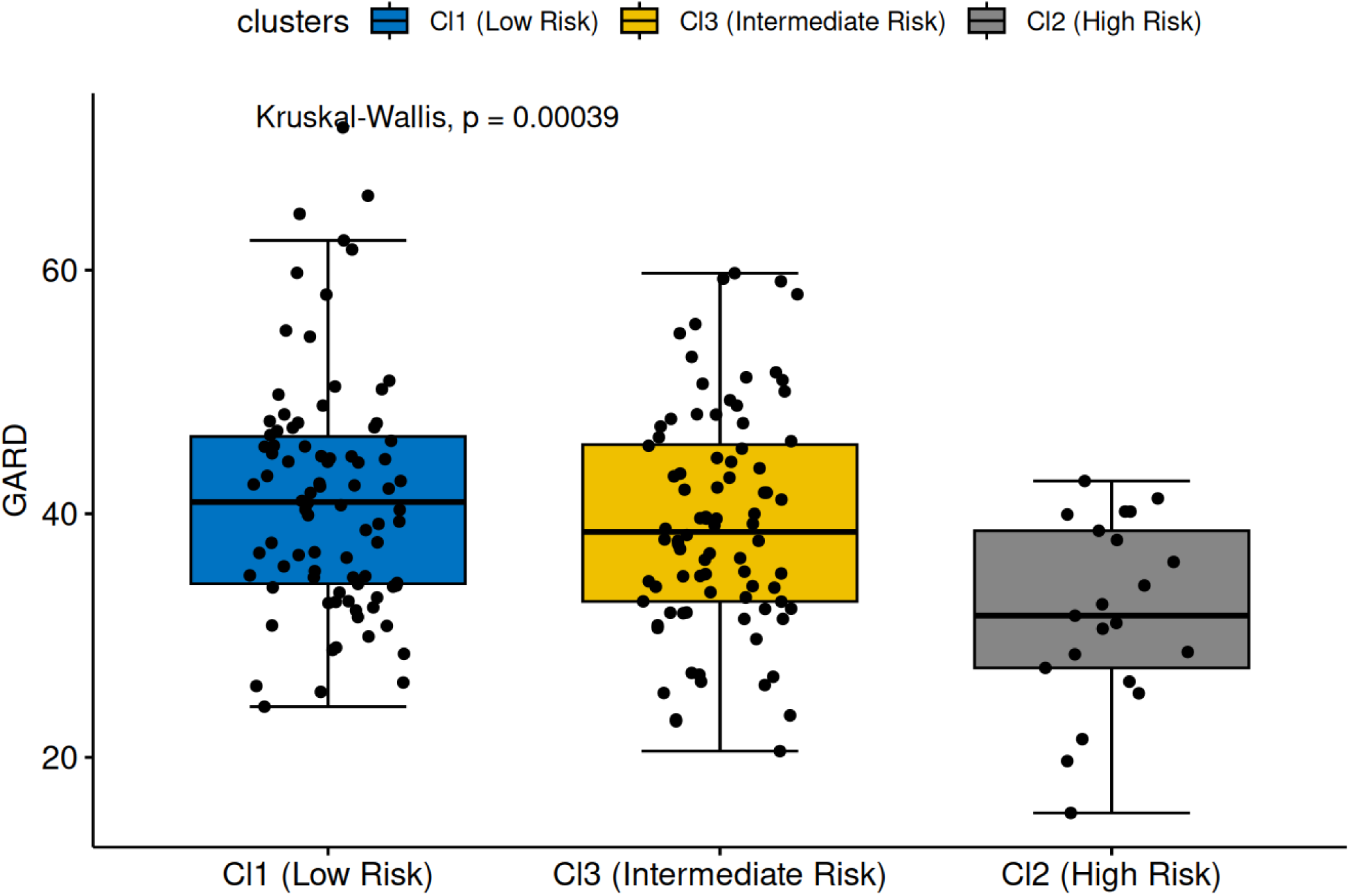
GARD values correlate with risk groups as calculated by gene expressions clusters. Boxplot of GARD values across the three defined cluster subtypes. GARD was calculated as described in the Methods for each patient.

### Calculating GARD thresholds

In order to find GARD cutpoints for our various *in silico* clinical trials, we performed cutpoint analyses with varying endpoints. First, to optimize outcome, we searched for the GARD threshold which optimally separated the groups by OS. In **Supplemental Figure 4**, we find a cutpoint of GARD 41.97 (42) optimally separates the groups.

**Supplemental Figure 4.**
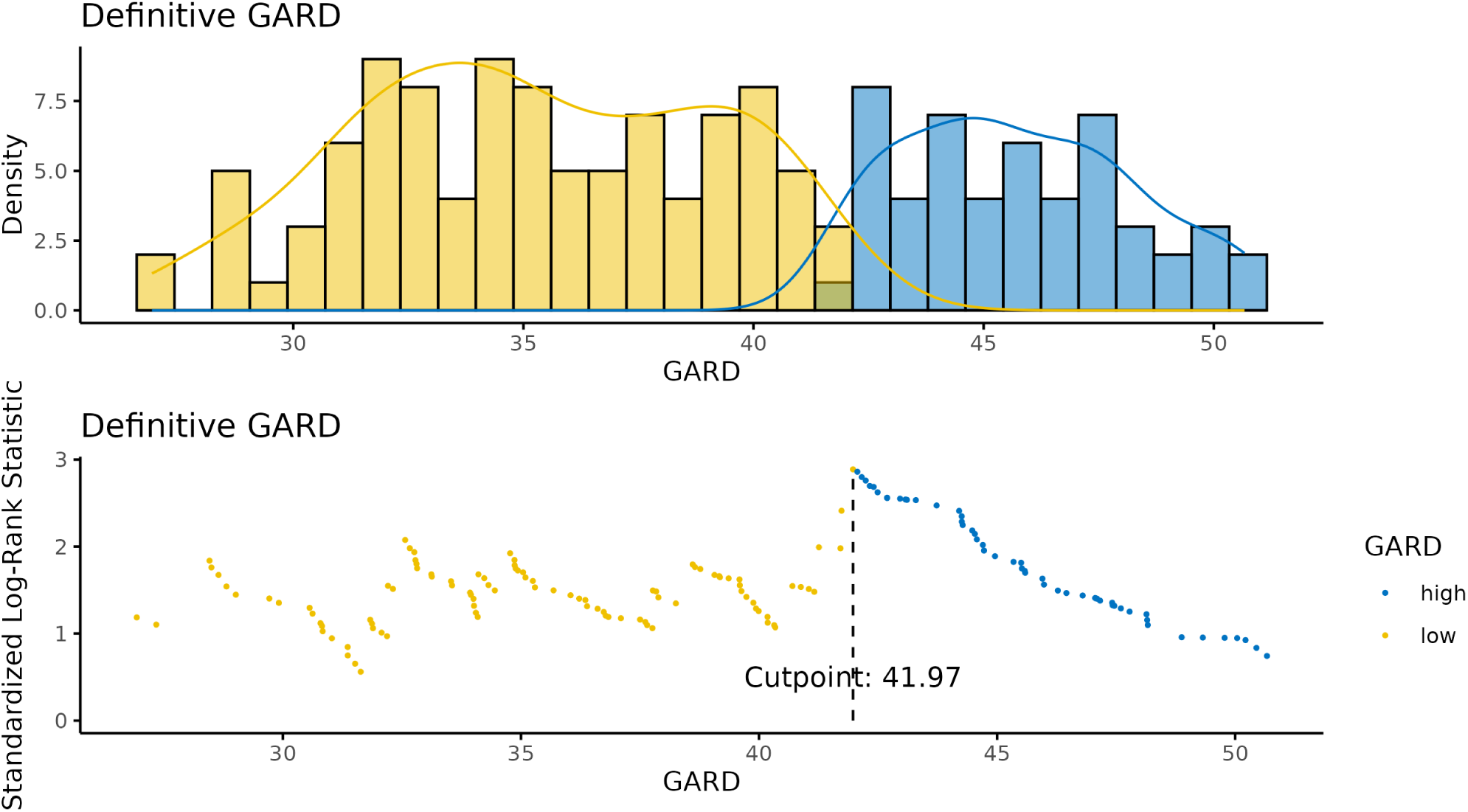
Identification of an optimal GARD threshold for outcome using survminer package and chisq statistic optimization.

We subsequently asked which GARD cutpoint would, at the population level, yield equipoise to the SOC, 70 Gy treatment. In **Supplemental Figure 5**, we find that a GARD cutpoint of 32 yields 97% OS at 3 years.

**Supplemental Figure 5.**
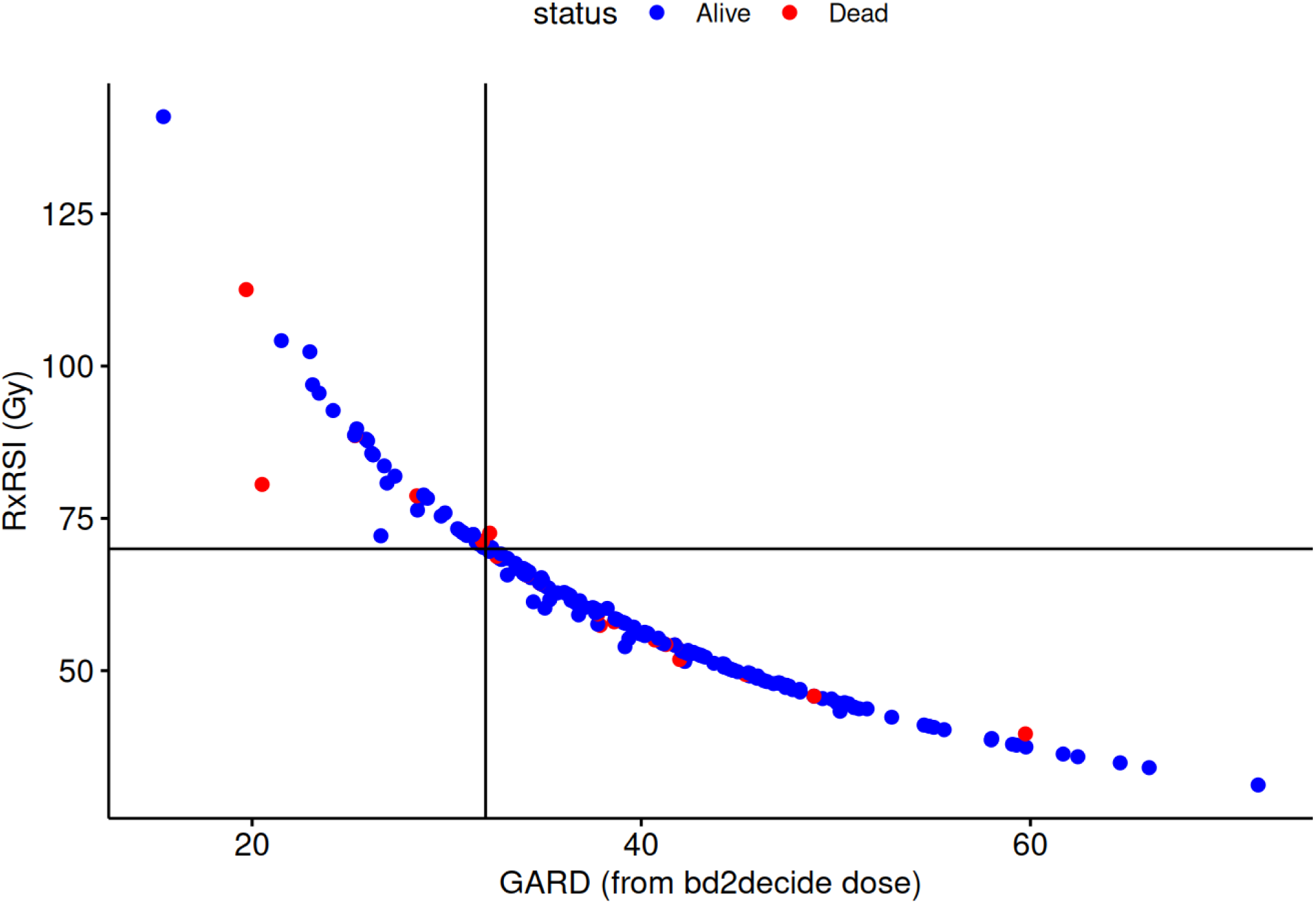
Identification of a GARD threshold for outcome to match SOC.

